# Violent offending in severe mental illness: the role of psychiatric comorbidity and crime type - insights from the first nationwide Norwegian registry linkage

**DOI:** 10.64898/2026.07.10.26357737

**Authors:** Martin Tesli, Seena Fazel, Lars Johan Hauge, Natalia Tesli, Stener Nerland, Marianne Riksheim Stavseth, Anne Bukten, Laoura Ziaka, Søren Esben Rytter Heilskov, Unn Kristin Haukvik, Anne Reneflot, Torbjørn Skardhamar, Christine Friestad, Jaroslav Rokicki

## Abstract

**Background:** Individuals with severe mental illness (SMI), including schizophrenia spectrum disorders (SSD) and bipolar disorder (BD), have been shown to have an elevated risk of violent perpetration. However, no population-wide study has systematically examined how this risk varies across psychiatric comorbidity patterns and specific violent crime types.

**Methods:** Using the first nationwide Norwegian registry linkage comprising mental health and crime data, we included 3,612,215 individuals aged 15-79 years living in Norway on Jan 1, 2008, and followed them until Dec 31, 2022. We estimated absolute and relative risks (RRs) of violent offending overall and by specific violent crimes among individuals with SSD and BD. To capture clinically relevant comorbidity patterns, we included substance use disorders (SUD), common personality disorders (PD), and hyperkinetic disorders (ADHD). RR models were adjusted first for sex and age, and subsequently for co-occurring mental disorders.

**Findings:** At the population level, individuals with SMI accounted for a minority of violent offenders (SSD: 8·7%; BD: 4·6%), whereas SUD was present among a substantially larger proportion (36·8%). Absolute risk of violent offending increased markedly with psychiatric comorbidity, from e.g., 5·0% among individuals with SSD alone to 43·9% for SSD combined with SUD and PD. Compared with the remaining general population, the RR of violent offending for SSD decreased from 6·58 (95% CI 6·4-6·8, adjusted for sex and age), to 2·0 (2·0-2·1) after further adjustment for other mental disorders. Similar attenuation patterns were observed across specific violent crime types, although varying in magnitude. In contrast to SMI, elevated risks associated with SUD remained substantial after full adjustment across most crime categories.

**Interpretation:** The association between SMI and violent offending is strongly influenced by psychiatric comorbidity, particularly SUD, and varies across crime types. Our findings underscore the importance of identifying and treating co-occurring mental disorders and substance use, both in the clinical management of SMI and in population-level violence prevention strategies.

**Funding:** Research Council of Norway (SAMRISK-2, No 341355). The funder had no role in study design, data collection, data analysis, data interpretation, writing of the report, or the decision to submit for publication.

## Introduction

Individuals with severe mental illness (SMI), including schizophrenia spectrum disorders (SSD) and bipolar disorder (BD), are not only experiencing significant personal distress but are also at increased risk of adverse outcomes such as violent offending. Although the majority of individuals with SMI will never commit a violent crime, the relative risk of violent offending is significantly elevated at group level^1^. The societal costs are substantial – a recent UK report estimated that violence related to SMI accounts for ∼15% of the total annual investment in adult mental health services^2^. Additionally, violent victimisation itself constitutes a significant public health issue^3^ contributing to excess morbidity and premature mortality globally, with disproportionate impact on socially and clinically vulnerable populations^4^.

The relationship between SMI and violence is complex and heterogenous. Risk varies substantially across diagnostic groups and is strongly influenced by co-occurring conditions such as substance use disorders (SUD), personality disorders (PD), and other clinical and social factors^5^. The public discourse on violence in SMI has often focused on rare but severe incidents, which may obscure the variation in risk largely observed across individuals and diagnostic profiles. A balanced account requires large, representative data sources that can distinguish between absolute and relative risk, identify clinically relevant high-risk subgroups, and avoid overgeneralising risk to all individuals with SMI.

Nordic administrative registers provide a unique opportunity to address these questions in population-wide cohorts^5^. Register-based studies from Sweden and Denmark have shown that people with mental disorders have increased rates of violent offending, with risk varying by age, sex, and diagnosis. Swedish studies have reported that individuals with mental disorders account for a minority of violent crimes but have higher individual-level risk than those without such disorders^6^. Danish registry studies have demonstrated similar patterns, with particularly high risks after onset of SSD, PD, and SUD^7–9^. Together, these studies provide strong evidence for an association between mental disorders and violent offending, while also indicating substantial heterogeneity across diagnostic groups.

However, registry-based studies have largely operationalised violence as a single composite outcome, without differentiating between specific categories of violent crime^3,6^. Disaggregating violence into distinct offence types may yield more granular insight into the nature and distribution of risk in the context of SMI. Although previous studies have reported elevated risks for specific offences – including arson, assault, and attempted homicide^10,11^ – these findings are constrained by small samples, selective focus on certain crime categories or diagnoses, and limited generalisability. To date, these questions have not been comprehensively examined in population-level registry data^3^.

Although co-occurring conditions such as SMI and SUD are consistently associated with increased risk of violent offending^12^, most previous studies have examined diagnostic categories in isolation, without accounting for the potential contribution of psychiatric comorbidity to violence risk^13^. Recent evidence from Nordic nationwide registry studies has shown that combinations of mental disorders are common^14^ and associated with an increased risk of adverse outcomes such as premature mortality^15^. To our knowledge, no previous study has systematically examined how combinations of mental disorders are associated with risk of violent offending in general and across specific crime categories.

To address these knowledge gaps, we used the first nationwide Norwegian linkage of population-based registry data on mental disorders and violent crime, covering the period 2008-2022. In the present study, we aim to 1) provide an overview of prevalence rates of violent offending in individuals with SMI compared to the general Norwegian population, 2) map the associations between SMIs and specific categories of violent offending, 3) estimate risk of total and specific violent offence taking into account combinations of SMI and other mental disorders and SUD, thereby identifying clinically relevant high-risk subgroups.

## Methods

### Data sources

The present study is part of the ForenPsych project (Clinical and sociodemographic risk factors of violence in mental disorders) which is based on a nationwide cohort derived from the Norwegian Population Registry and includes all individuals residing in Norway from 1960 onwards. The ForenPsych project aims to identify risk factors and characterise subgroups with increased vulnerability to violent offending among individuals with diagnosed mental disorders in Norway. ForenPsych has an approval from The Regional Committees for Medical and Health Research Ethics South-East (226893), as well as The Data Protection Impact Assessment (DPIA) at the Norwegian Institute of Public Health. We report this study in accordance with the REporting of studies Conducted using Observational Routinely-collected Data (RECORD) statement, an extension of STROBE for routinely collected health data^16^.

### Study population

The study population was identified from the Norwegian Population Registry and comprised all individuals aged 15-79 with a personal identification number (PIN) residing in Norway on Jan 1, 2008 (N=3,612,215), with follow-up period through Dec 31, 2022. A PIN is given to persons who are born in Norway, have a valid residence permit for at least six months or have officially moved to Norway. Demographic characteristics are presented in **Table 1**.

**Table 1:** Sex-specific prevalence of violent crime by diagnostic group. Data are retrieved from Norwegian national registries and include all individuals living in Norway who were aged 15-79 years on Jan 1, 2008, with follow-up to Dec 31, 2022. Mental disorders were identified from the Norwegian Patient Registry and violent crime convictions from Statistics Norway. Percentages in the violent crime columns are calculated using the sex-specific diagnostic-group denominator shown in the adjacent N column. The violent-crime cohort percentage is the proportion of all individuals with violent crime in the corresponding sex-specific population accounted for by each diagnostic group. Diagnostic groups are not mutually exclusive. ICD-10=International Classification of Diseases, 10th revision.

| Diagnostic group | Female |  |  | Male |  |  |
| --- | --- | --- | --- | --- | --- | --- |
|  | N | Violent crime n (%) | % of violent crime cohort | N | Violent crime n (%) | % of violent crime cohort |
| **Overall population** | **1,792,699** | **9,725 (0.5%)** | **100.0** | **1,819,516** | **53,805 (3.0%)** | **100.0** |
| Schizophrenia spectrum disorders | 19,367 | 1,099 (5.7%) | 11.3 | 22,718 | 4,431 (19.5%) | 8.2 |
| Bipolar disorder | 25,224 | 736 (2.9%) | 7.6 | 18,112 | 2,191 (12.1%) | 4.1 |
| Substance use disorders | 56,003 | 3,501 (6.3%) | 36.0 | 102,605 | 19,855 (19.4%) | 36.9 |
| Personality disorders | 15,275 | 1,249 (8.2%) | 12.8 | 9,264 | 3,020 (32.6%) | 5.6 |
| Hyperkinetic disorders | 27,439 | 1,364 (5.0%) | 14.0 | 31,048 | 6,852 (22.1%) | 12.7 |

### Mental disorders and violent crime types

Diagnoses of mental disorders were identified from registrations in the Norwegian Patient Registry (NPR) according to the International Classification of Diseases, 10th revision (ICD-10)^17^. SMIs included the following diagnoses: schizophrenia, schizotypal, and delusional disorder (F20-F29); “SSD”) or a diagnosis of manic episode and bipolar affective disorder (F30-F31; “BD”). We also included diagnostic categories previously identified as related to violent offending^18–20^: a) Mental and Behavioural disorders due to psychoactive substance use (F10-F19; “SUD”), (b) common personality disorders (“PD”): Paranoid personality disorder (F60.0), Schizoid personality disorder (F60.1), Dissocial personality disorder (F60.2), Emotionally unstable (Borderline) personality disorder (F60.3), Histrionic personality disorder (F60.4), and (c) Behavioural and emotional disorders with onset usually occurring in childhood and adolescence: Hyperkinetic disorders (F90; “ADHD”). Diagnostic groups were not mutually exclusive. The onset of mental disorder was operationalised as the first registered contact with the health services for the diagnosis of interest within the period of observation. The date of contact with health care services was recorded as the month and year in which the contact occurred.

Violent offending was defined as a registered charge for an offence classified under Statistics Norway’s “Violence and maltreatment” category, with legal decision year between 2008 and 2022, excluding charges ending in acquittal. We restricted the main analyses to convicted offences; individuals who were acquitted were classified as non-cases. In sensitivity analyses, we repeated all analyses using violent charges (regardless of conviction) as the outcome. For offence-specific analyses, we used the underlying offence codes in this category to define distinct violent crime types (**Supplementary Table 2**). Because NPR data were available from 2008 onwards, only individuals with decisions on violent offences registered after Jan 1, 2008, were included in the analyses to match the NPR data. As such, decisions on criminal offences registered before 2008 were not taken into account in the statistical analyses.

### Statistical analyses

### Demographics

For each diagnostic group, we calculated the proportion of individuals registered with at least one violent offence, and each group’s contribution to the overall violent-offending cohort. We further estimated age-specific counts of unique individuals convicted of a violent offence and visualised these as age-crime curves according to age at registered offence. All analyses were stratified by sex.

### Main analyses

We conducted two sets of analyses. First, we examined the associations between individual and co-occurring mental disorders and risk of any violent crime. Second, we examined the association between mental disorders and specific violent crime types.

We used UpSet plots^21^ to visualise the frequency of combinations of registrations of SSD, BD, SUD, PD, and ADHD throughout the observation period. For each diagnostic category and diagnostic combination, we calculated the absolute risk of any violent crime during follow-up as the number of individuals with at least one violent crime conviction divided by the total number of individuals in the corresponding diagnostic subgroup.

We estimated relative risks of ever being convicted for violent offense as binary outcome for each mental disorder using Poisson regressions with robust standard errors^22^. The comparison groups comprised all individuals without the disorder of interest. For each disorder, we fitted two regression models with any violent crime conviction during follow-up as the outcome. The first model (model 1) adjusted for sex and age. The second model (model 2) additionally adjusted for the other mental disorders included in the analysis, thereby estimating the association between the disorder of interest and violent crime after accounting for measured psychiatric comorbidity.

We then examined specific violent crime types. For each violent crime type, we calculated the proportion of convicted individuals with each mental disorder, enabling comparison of diagnostic profiles across offence categories. We further estimated relative risks with Poisson regressions for each specific violent crime type in relation to each mental disorder, again comparing individuals with the diagnosis of interest with the remaining general population. As for the overall violence analyses, models were first adjusted for sex and age and then additionally adjusted for the other mental disorders. These analyses were used to assess whether associations between mental disorders and violent offending varied by offence type and whether risk estimates were attenuated after adjustment for psychiatric comorbidity. The distribution of offence types was described for the total population and by combinations of diagnostic groups using pie charts (excluding crime types with N<50 individuals). As a sensitivity analysis, we repeated all analyses using charges rather than convictions as the outcome, to assess whether the findings were robust to the inclusion of violent offences that did not result in conviction. The analyses were based exclusively on routinely collected data from nationwide administrative registries, and no imputation or other procedures for handling missing data were required.

## Results

### Demographics

The study population comprised 3,612,215 individuals, including 1,792,699 females and 1,819,516 males. Overall, 9,725 females (0·5%) and 53,805 males (3·0%) had a registered violent crime. The age distribution of violent convictions followed the typical age–crime curve. (**Supplementary Figure 1**). Across diagnostic groups, the largest contribution to the violent-crime cohort was observed among individuals with SUD. Among females with SUD, 6·3% had a registered violent crime, accounting for 36·0% of all females with violent crime. Among males, the corresponding figures were 19·4% with violent crime, accounting for 36·9% of all males with violent crime (**Figure 1**). ADHD was the second-largest contributor among females (14·0%) and males (12·7%), whereas SSD accounted for 11·3% of females and 8·2% of males with violent crime, and BD for 7·6% and 4·1% of females and males, respectively.

**Figure 1.**
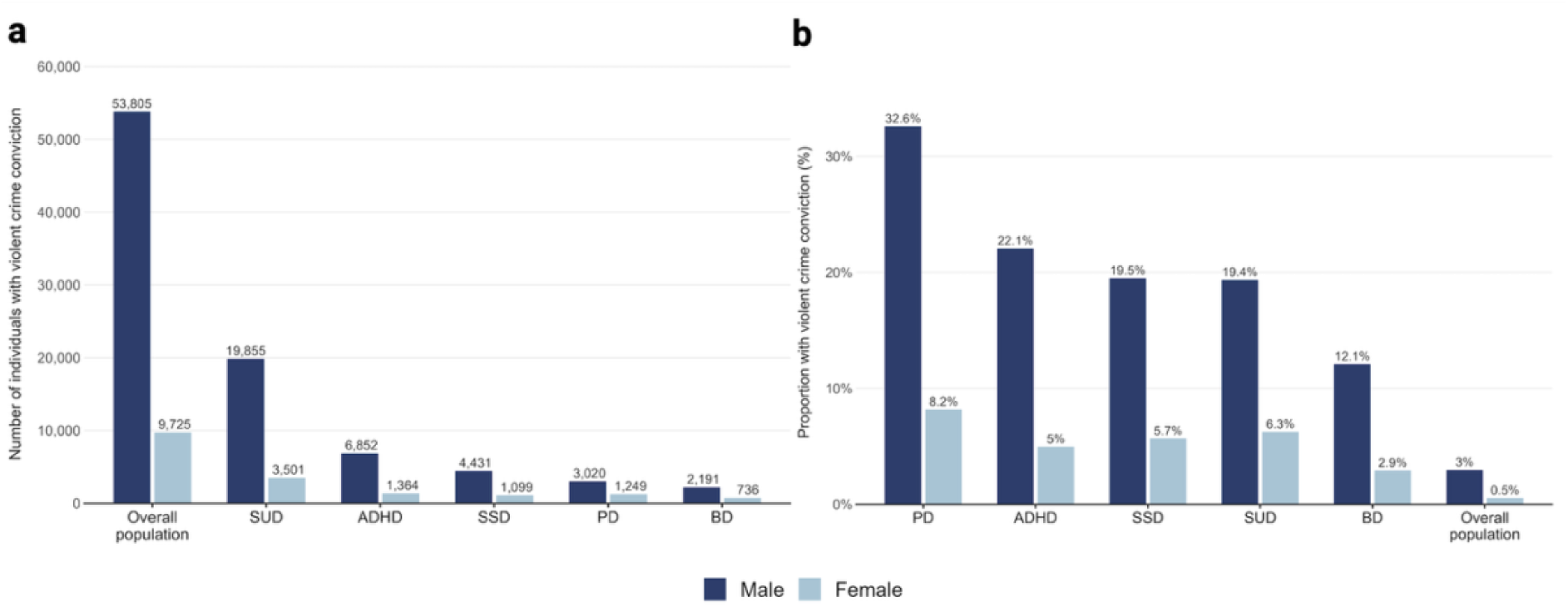
**a**: Number of individuals convicted for violent crime, by diagnostic category and sex. **b.** Proportion of individuals within each diagnostic category convicted for violent crime, by sex. The population comprises all Norwegian residents aged 15-79 Jan 1 2008, followed to Dec 31, 2022. All diagnostic categories are classified according to ICD-10. ADHD=Hyperkinetic disorder (F90). BD=Bipolar disorder (F30-F31). PD=Common personality disorders (F60.0, F60.1, F60.2, F60.3, F60.4). SSD=Schizophrenia spectrum disorders (F20-F29). SUD=Substance use disorders (F10-F19).

Among females, the highest within-diagnostic-group proportion with violent crime was observed for PD (8·2%), followed by SUD (6·3%), SSD (5·7%), ADHD (5·0%), and BD (2·9%). Among males, the highest within-diagnostic-group proportion with violent crime was also observed for PD (32·6%), followed by ADHD (22·1%), SSD (19·5%), SUD (19·4%), and BD (12·1%) (**Figure 1**). Please see **Table 1** for further demographic information.

### Combinations of mental disorders and absolute risk of overall violence

The absolute risk of violent offending during the 15-year observation period varied substantially by sex and diagnostic comorbidity. Among individuals with SSD alone, the absolute risk was 7·8% in males and 2·2% in females, compared with 3·0% and 0·5%, respectively, in the general population. The risk increased markedly in the presence of comorbid SUD, reaching 32·2% in males and 13·0% in females with SSD and SUD. The highest risks were observed among individuals with SSD combined with both SUD, ADHD and severe PD, among whom 71·5% of males and 36·9% of females had a registered violent crime during follow-up. A similar, although less pronounced pattern, was observed for BD, with higher absolute risks among individuals with comorbid SUD and other mental disorders than among those with BD alone. The distribution of diagnostic combinations and corresponding absolute risks is shown in the UpSet plot in **Figure 2**.

**Figure 2.**
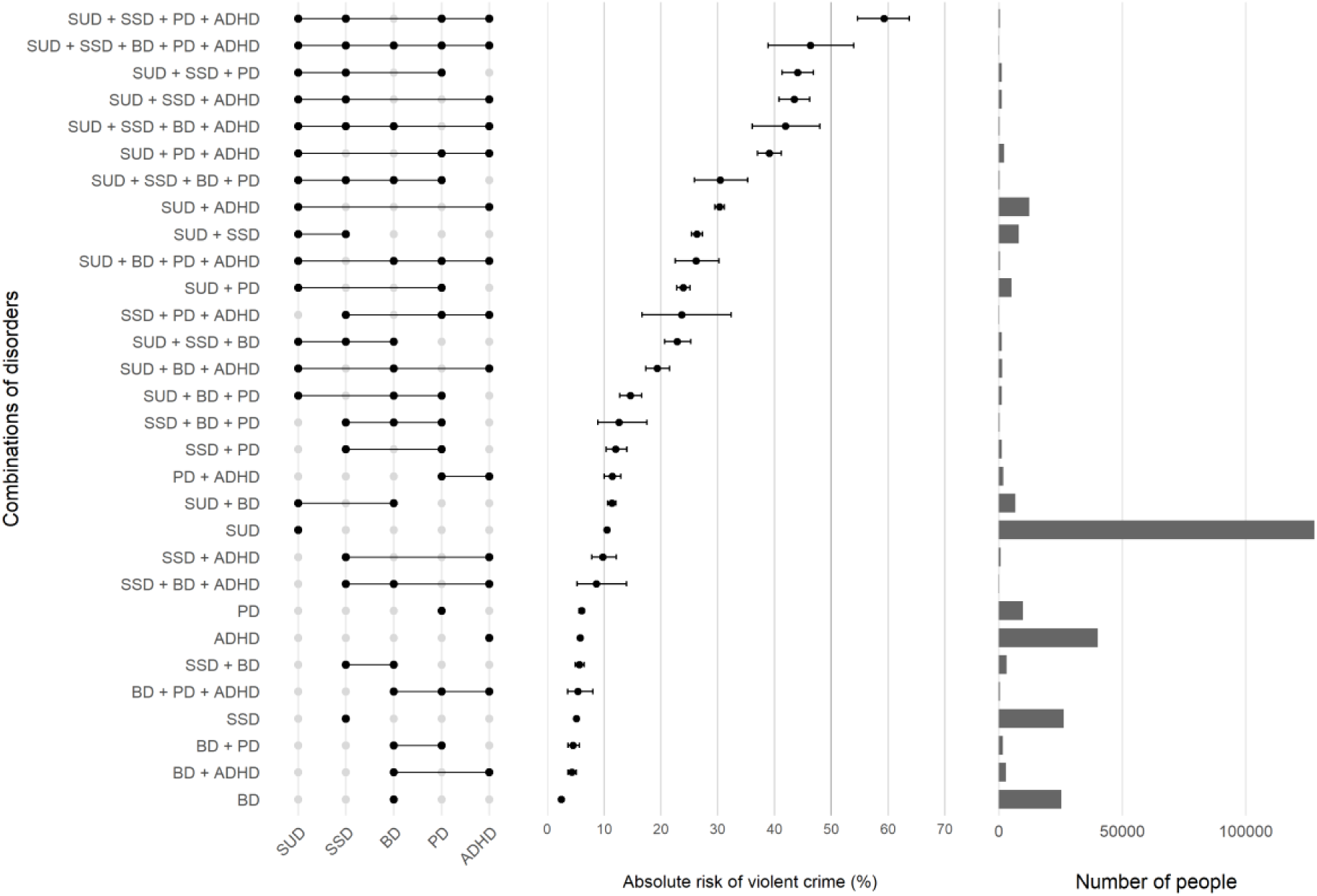
UpSet plot of absolute risk of conviction of violent offending by group of combinations of mental disorders in the entire Norwegian population aged 15-79 Jan 1, 2008, followed to Dec 31, 2022. Number of individuals, marked by horizontal bars for each diagnostic group and group combination are displayed in the right panel. All diagnostic categories are classified according to ICD-10. ADHD=Hyperkinetic disorder (F90). BD=Bipolar disorder (F30-F31). PD=Common personality disorders (F60.0, F60.1, F60.2, F60.3, F60.4). SSD=Schizophrenia spectrum disorders (F20-F29). SUD=Substance use disorders (F10-F19).

### Combinations of mental disorders and relative risk of overall violence

In models adjusted for sex and age, SSD was associated with an increased risk of violent offending compared with the remaining general population (RR 6·6, 95% CI 6·4-6·8; **Supplementary Table 1, model 1**). This association was substantially attenuated after further adjustment for other mental disorders, including SUD, PD, ADHD, and BD, but remained elevated (RR 2·0, 95% CI 2·0-2·1; **Supplementary Table 1, model 2**). A similar pattern was observed for BD: the sex– and age-adjusted RR was 3·9 (95% CI 3·7-4·0), decreasing to 1·2 (1·2-1·3) after adjustment for other mental disorders. By contrast, the association between SUD and violent offending remained strong after adjustment for psychiatric comorbidity. The RR for SUD decreased from 9·2 (95% CI 9·0-9·3) in the sex– and age-adjusted model to 6·9 (6·8-7·0) in the fully adjusted model.

### Risk of specific violent crime types in mental disorders

Assault was the most common violent crime type among people convicted for any violence in the general population, among both females and males, followed by reckless behaviour and stalking. More severe offences, including homicide and attempted homicide, were rare. Across all violent crime categories, offence rates were higher in males than in females (**Supplementary Table 2, Supplementary Figure 2**).

The proportion of individuals with a registered SMI (SSD and/or BD) was in general low across all violent crime types (**Supplementary Figure 3a**). However, these proportions increased significantly when including any mental disorder (SSD, BD, SUD, ADHD, PD) instead of only SMI (SSD, BD). The proportion in the total diagnostic group (any mental disorder) was highest for offences such as assaults or threats against public servants and robbery, and lowest for unintentional bodily harm and manslaughter (**Supplementary Figure 3b**). When examined by diagnostic category, SUD accounted for the largest share across most violent crime types, with the exception of unintentional bodily harm and manslaughter. A broadly similar, although less pronounced, pattern was observed for SSD.

Relative risk estimates also varied by offence type and were generally attenuated after adjustment for other mental disorders. For example, the sex– and age-adjusted RR for assault among individuals with SSD was 5·5 (95% CI 5·3-5·7), decreasing to 1·7 (1·6-1·8) after adjustment for other mental disorders. For manslaughter, the corresponding RRs were 1·7 (95% CI 0·8-3·5) and 0·7 (0·3-1·5), respectively. In contrast to SSD and BD, elevated risks associated with SUD remained substantial after full adjustment across most specific violent crime types. For the most severe albeit rare types of violent crime such as murder and attempted murder, RR remained high also after adjustment for age, sex and all other mental disorders for SSD and SUD (**Supplementary Table 3a** and **Supplementary Table 3b**).

When patterns of diagnostic comorbidity were examined separately for each violent crime type, SUD was the dominant contributor to most diagnostic combinations across offence categories. This pattern was less evident for unintentional bodily harm and manslaughter, for which psychiatric comorbidity appeared less prominent. For distribution of specific violent crime types across combinations of mental disorders and SUD, please see pie charts in **Figure 3**.

**Figure 3.**
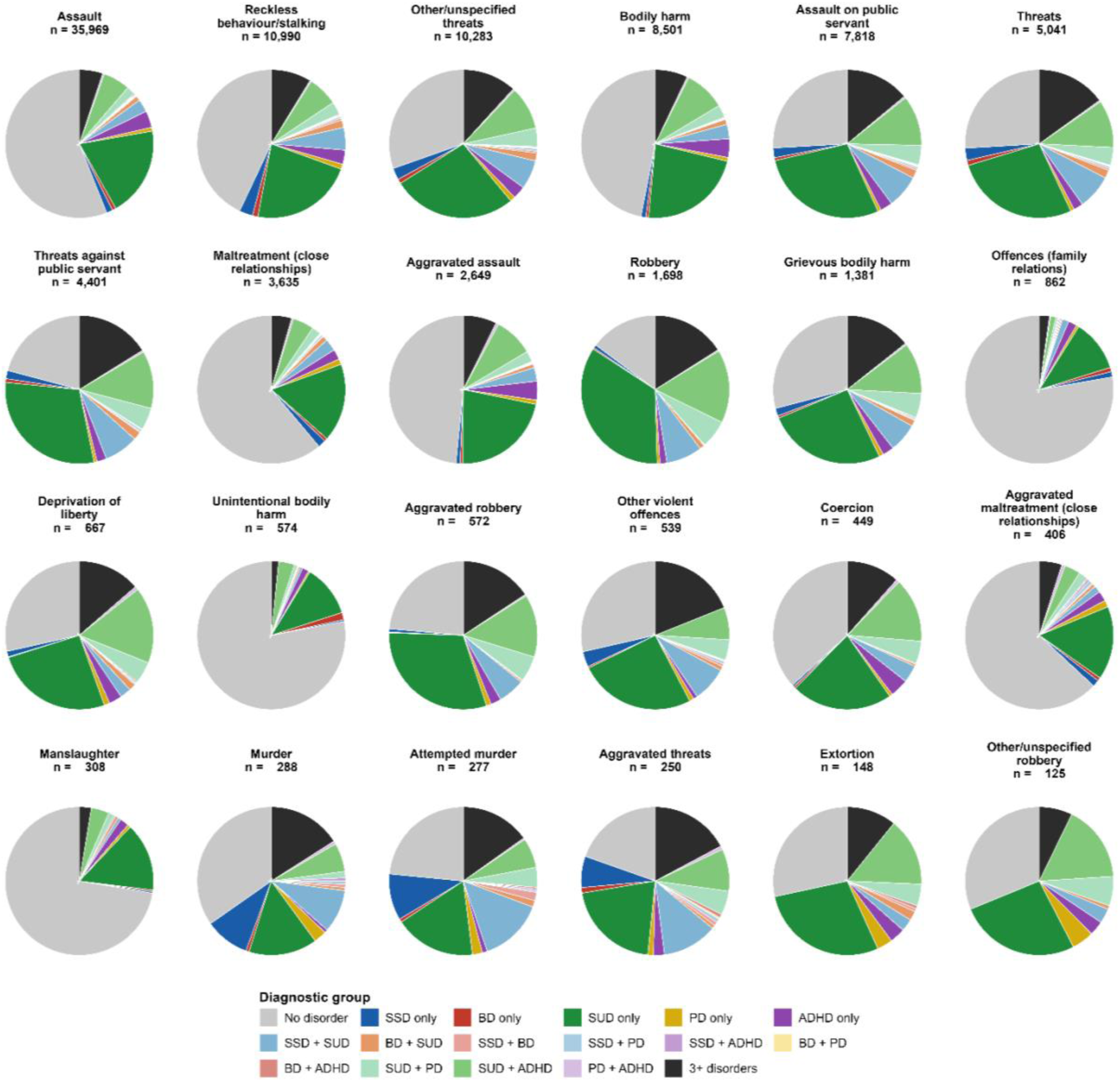
Proportion of specific violent crime types across combinations of mental disorders in the entire Norwegian population aged 15-79 Jan 1, 2008, followed to Dec 31, 2022. All diagnostic categories are classified according to ICD-10. ADHD=Hyperkinetic disorder (F90). BD=Bipolar disorder (F30-F31). PD=Common personality disorders (F60.0, F60.1, F60.2, F60.3, F60.4). SSD=Schizophrenia spectrum disorders (F20-F29). SUD=Substance use disorders (F10-F19).

In the sensitivity analyses, the findings were largely unchanged when charges were used instead of convictions as the outcome.

## Discussion

In this nationwide registry-linkage study of 3,612,215 individuals followed for 15 years, we found that SMI was associated with increased risk of violent offending, but risks varied according to psychiatric comorbidity and offence type. Individuals with SSD and BD accounted for a minority of violent crime at the population level, whereas SUD accounted for a substantially larger proportion. Absolute risks were markedly elevated among individuals with co-occurring mental disorders, while relative risk estimates for SSD and BD were substantially attenuated after adjustment for psychiatric comorbidity. In contrast, the association between SUD and violent offending remained strong after adjustment for other mental disorders. These findings indicate that violence risk in people with SMI is concentrated within clinically important subgroups characterised by mental multimorbidity, rather than distributed uniformly across diagnostic categories.

Our results are consistent with previous population-based studies from Sweden and Denmark showing increased rates of violent offending among people with psychiatric disorders, particularly among those with SSD, PD, and SUD^1,3^. However, the present study extends this evidence in several important ways. First, we examined diagnostic overlap explicitly, rather than treating psychiatric diagnoses as isolated or mutually exclusive categories. Second, we disaggregated violent crime into specific offence types, demonstrating that associations between mental disorders and violence vary across offence categories and are not adequately captured by a single composite violence outcome. Third, the first nationwide Norwegian linkage of mental health and crime registries allowed us to extend existing Scandinavian evidence on the association between SMI and violence using a contemporary national cohort covering a more recent period than comparable Nordic studies^3,23^.

Although relative risks for violent offending were increased among individuals with SMI, absolute risks remained modest for those with SSD or BD alone, particularly among women. This distinction between absolute and relative risk is clinically and epidemiologically important. Relative risks quantify group-level associations, but can overstate perceived individual risk when base rates are low. Conversely, absolute risks are more directly relevant for clinical communication and service planning. The current findings therefore support a balanced interpretation: SMI is associated with increased violence risk at the population level, but violent offending remains uncommon for most individuals with these disorders.

The strong attenuation of risk estimates after adjustment for co-occurring mental disorders suggests that psychiatric comorbidity accounts for an important part of the association between SMI and violent offending. For SSD, the relative risk decreased from 6·6 after adjustment for sex and age to 2·0 after further adjustment for other mental disorders, while the corresponding estimate for BD decreased from 3·9 to 1·2. These findings indicate that violence risk in SMI is strongly shaped by diagnostic context and comorbidity patterns rather than by isolated diagnoses alone. In clinical practice, individuals often present with complex patterns of diagnostic comorbidity. SUD, personality pathology, ADHD, previous adversity, social marginalisation, and impaired treatment engagement may cluster with SMI and shape risk trajectories. By quantifying overlapping diagnostic patterns, our study identifies clinically relevant subgroups for whom preventive interventions may be particularly important.

SUD emerged as the most consistent and substantial correlate of violent offending. This diagnostic category contributed the largest absolute number of individuals with violent crime in both sexes and remained strongly associated with violence after adjustment for other mental disorders. This pattern was observed not only for overall violence but also across most specific violent crime types. These findings align with a large body of evidence identifying substance use as a key modifiable risk factor for violence in psychiatric and general populations^12,24^. They also have direct implications for services. Integrated treatment for substance use should be considered a core component of violence prevention in people with SMI, rather than an adjunctive or secondary concern. Early identification and treatment of substance use, combined with continuity of care, relapse prevention, and assertive follow-up after acute episodes or service disengagement, may be among the most actionable clinical strategies suggested by our findings.

The offence-specific analyses showed that psychiatric associations varied across violent crime types. The proportion of offenders with any registered mental disorder was highest for offences such as attacks or threats against public servants and robbery, and lowest for unintentional bodily harm and manslaughter. Relative risk estimates also varied by offence category and were attenuated after comorbidity adjustment, particularly for SMI, with a notable exception of SSD and homicide/attempted homicide. This heterogeneity has implications for research and policy. Studies that combine all violent offences into a single outcome may obscure clinically and criminologically meaningful differences between offence types. Less severe but more frequent offences, such as assault and threats, are likely to drive most population-level associations, whereas rare severe offences generate imprecise estimates and should be interpreted cautiously. Future work should examine whether offence-specific patterns reflect differences in situational context, victim-offender relationships, intoxication, acute symptoms, social adversity, or criminal justice detection.

This study has several strengths. It used a nationwide cohort with long follow-up, minimising selection bias and enabling precise estimation for relatively uncommon diagnostic groups and offence categories. Linkage of health and crime registers allowed us to examine mental disorders and violent convictions at the population level, rather than relying on clinical samples, prison samples, or self-report. The large sample also enabled sex-stratified descriptive analyses and assessment of subgroups of co-occurring diagnostic categories. Moreover, the current study comprised more recent data than previously published comparable reports^1,3,23^. Finally, the use of offence-specific outcomes provides a more granular account of violence than has typically been available in previous registry-based studies.

### Limitations

Several limitations should be considered. First, violent offending was defined using convictions. This captures legally processed violence, but not all violent behaviour – as offences that are not reported, investigated, prosecuted, or resulting in conviction are not included. However, sensitivity analyses using charged rather than convicted individuals yielded largely similar findings (results not shown). Detection and prosecution may also vary by offence type, sex, socioeconomic position, substance use, and psychiatric status, which could influence observed associations. Second, registry diagnoses reflect contact with specialist health services and recorded clinical diagnoses. Individuals with untreated, undiagnosed, or primary-care-managed disorders may have been misclassified as unexposed. This limitation is particularly relevant for SUD^25^ and PD^26^, which are often underdiagnosed.

Third, diagnostic categories were treated as time-fixed indicators of registered diagnoses during the observation period. This approach is suitable for describing population-level associations and comorbidity patterns, but does not establish the temporal ordering of disorder onset, comorbidity, and violent offending for each individual. Future analyses should model time-varying exposures and post-diagnosis risk trajectories. Fourth, although the models adjusted for sex, age, and other mental disorders, they did not adjust for e.g., socioeconomic position or common genetic risk. These factors may confound or mediate associations between mental disorders and violence. The fully adjusted estimates should therefore not be interpreted as causal effects of diagnostic categories.

Fifth, some specific violent crime types were rare, particularly severe offences such as homicide and manslaughter. Estimates for these outcomes are therefore less precise and should be interpreted with caution, even in a nationwide cohort. Sixth, the study was conducted in Norway, a high-income country with universal health care, comparatively low rates of violent crime, and a specific legal and welfare context. The absolute risks and population-attributable patterns may not generalise directly to countries with different health-care systems, criminal justice practices, firearm availability, substance use patterns, or social inequalities. Finally, we focused on diagnostic categories and convictions and could not examine clinical state at the time of the offence, including active psychotic symptoms, mood episodes, intoxication, treatment adherence, medication exposure, or service contact immediately before offending.

### Conclusion

This nationwide Norwegian registry-linkage study shows that SMI is associated with increased risk of violent offending, but that the association is strongly shaped by psychiatric comorbidity and varies across offence types. The highest absolute risks were observed among individuals with several co-occurring mental disorders, and associations for SSD and BD were substantially attenuated after adjustment for other mental disorders. SUD showed the most robust and consistent association with violent offending across offence categories. These findings support a shift from diagnosis-based interpretations of violence risk towards a comorbidity-informed approach, with particular emphasis on integrated treatment of substance use, early identification of multimorbidity, and targeted clinical follow-up for high-risk subgroups. Such an approach may improve risk stratification and support more targeted violence prevention strategies in psychiatric services.

## Ethics Statement

ForenPsych has an approval from The Regional Committees for Medical and Health Research Ethics South-East (226893), as well as The Data Protection Impact Assessment (DPIA) at the Norwegian Institute of Public Health. All methods were performed in accordance with relevant guidelines and regulations including the Declarations of Helsinki.

## Data Availability Statement

The data that support the findings of this study are available from the Norwegian Health Data Authority and Statistics Norway, but restrictions apply to the availability of these data, which were used under license for the current study. Data are, however, available from the corresponding author, MT, upon reasonable request and with permission of the Norwegian Health Data Authority and Statistics Norway.

