## Supplementary material for "Violent offending in severe mental illness: the role of psychiatric comorbidity and crime type - insights from the first nationwide Norwegian registry linkage"

**Supplementary Figure 1. Age–crime curve: number of individuals with at least one violent crime conviction, by age at criminal offence and sex.**

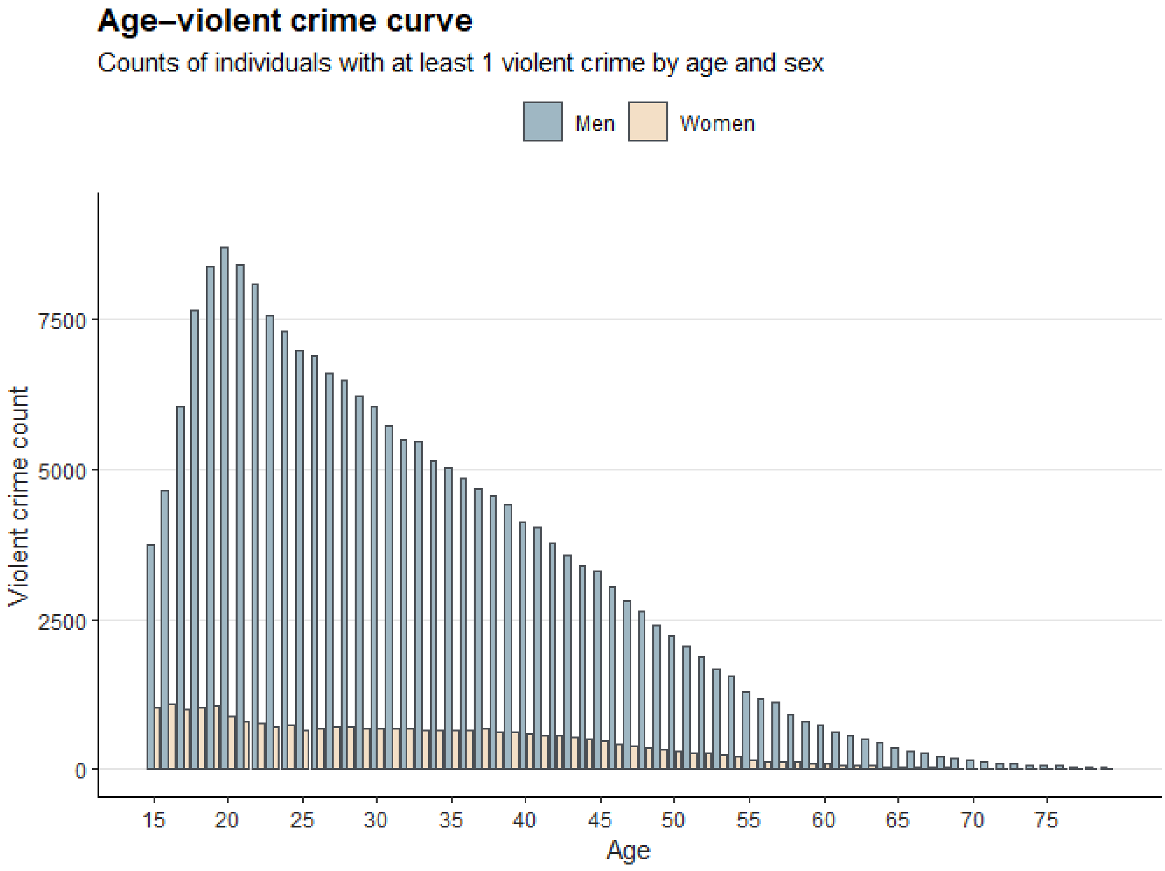

**Supplementary Table 1. Relative risk of any violent crime conviction associated with each mental disorder, before and after adjustment for psychiatric comorbidity.**

| | Both models | | Model 1: sex + age adjusted | | Model 2: fully adjusted | |
|---|---|---|---|---|---|---|
| **Diagnosis** | **N** | **Violent crime convictions n** | **RR (95% CI)** | **p** | **RR (95% CI)** | **p** |
| SSD | 42,085 | 5,530 | 6·58 (6·42–6·75) | <0·001 | 2·03 (1·98–2·09) | <0·001 |
| BD | 43,336 | 2,927 | 3·87 (3·73–4·01) | <0·001 | 1·21 (1·16–1·25) | <0·001 |
| SUD | 158,608 | 23,356 | 9·18 (9·04–9·32) | <0·001 | 6·90 (6·77–7·02) | <0·001 |
| PD | 24,539 | 4,269 | 8·35 (8·13–8·59) | <0·001 | 1·96 (1·90–2·02) | <0·001 |
| ADHD | 58,487 | 8,216 | 4·96 (4·85–5·07) | <0·001 | 1·83 (1·79–1·88) | <0·001 |

**Supplementary table 1.** *Poisson regression with robust standard errors. Outcome: any violent crime conviction (binary yes/no). Reference group: remaining general population (all individuals without the disorder of interest). Model 1: adjusted for sex and age. Model 2: additionally adjusted for all other mental disorders (SSD, BD, SUD, PD, ADHD). RR = relative risk; CI = confidence interval. SSD = schizophrenia spectrum disorders (F20–F29); BD = bipolar disorder (F30–F31); SUD = substance use disorders (F10–F19); PD = common personality disorders (F60·0, F60·1, F60·2, F60·3, F60·4); ADHD = hyperkinetic disorders (F90).*

### Supplementary Table 2: Frequency of specific violent crime types among convicted individuals, by sex.

| | Female (N = 9,725) | Male (N = 53,805) | Total (N = 63,530) |
|---|---|---|---|
| **Violent crime type** | **Female n (%)** | **Male n (%)** | **Total n (%)** |
| Assault | 5,497 (56·5%) | 30,472 (56·6%) | 35,969 (56·6%) |
| Reckless behaviour/stalking | 1,527 (15·7%) | 9,463 (17·6%) | 10,990 (17·3%) |
| Other/unspecified threats | 844 (8·7%) | 9,439 (17·5%) | 10,283 (16·2%) |
| Bodily harm | 499 (5·1%) | 8,002 (14·9%) | 8,501 (13·4%) |
| Assault on public servant | 1,676 (17·2%) | 6,142 (11·4%) | 7,818 (12·3%) |
| Threats | 513 (5·3%) | 4,528 (8·4%) | 5,041 (7·9%) |
| Threats against public servant | 322 (3·3%) | 4,079 (7·6%) | 4,401 (6·9%) |
| Maltreatment (close relationships) | 552 (5·7%) | 3,083 (5·7%) | 3,635 (5·7%) |
| Aggravated assault | 260 (2·7%) | 2,389 (4·4%) | 2,649 (4·2%) |
| Robbery | 113 (1·2%) | 1,585 (2·9%) | 1,698 (2·7%) |
| Grievous bodily harm | 146 (1·5%) | 1,235 (2·3%) | 1,381 (2·2%) |
| Offences (family relations) | 175 (1·8%) | 687 (1·3%) | 862 (1·4%) |
| Deprivation of liberty | 58 (0·6%) | 609 (1·1%) | 667 (1%) |
| Unintentional bodily harm | 94 (1%) | 480 (0·9%) | 574 (0·9%) |
| Aggravated robbery | 35 (0·4%) | 537 (1%) | 572 (0·9%) |
| Other violent offences | 129 (1·3%) | 410 (0·8%) | 539 (0·8%) |
| Coercion | 37 (0·4%) | 412 (0·8%) | 449 (0·7%) |
| Aggravated maltreatment (close relationships) | 64 (0·7%) | 342 (0·6%) | 406 (0·6%) |
| Manslaughter | 37 (0·4%) | 271 (0·5%) | 308 (0·5%) |
| Murder | 29 (0·3%) | 259 (0·5%) | 288 (0·5%) |
| Attempted murder | 39 (0·4%) | 238 (0·4%) | 277 (0·4%) |
| Aggravated threats | 23 (0·2%) | 227 (0·4%) | 250 (0·4%) |
| Extortion | 8 (0·1%) | 140 (0·3%) | 148 (0·2%) |
| Other/unspecified robbery | 7 (0·1%) | 118 (0·2%) | 125 (0·2%) |
| Human trafficking | 9 (0·1%) | 32 (0·1%) | 41 (0·1%) |
| Terrorism-related | NA | 28 (0·1%) | 30 (0%) |

*Crime types ordered by total count (most to least frequent). % = proportion of all convicted individuals in the sex-specific population with a conviction of that crime type. Individuals may have convictions for more than one crime type. Data from Norwegian national mandatory registries; all individuals aged 15-79 residing in Norway on Jan 1, 2008, followed to Dec 31, 2022. NA=too few observations in group.*

**Supplementary Figure 2. Number of individuals convicted of each violent crime type, by sex.**

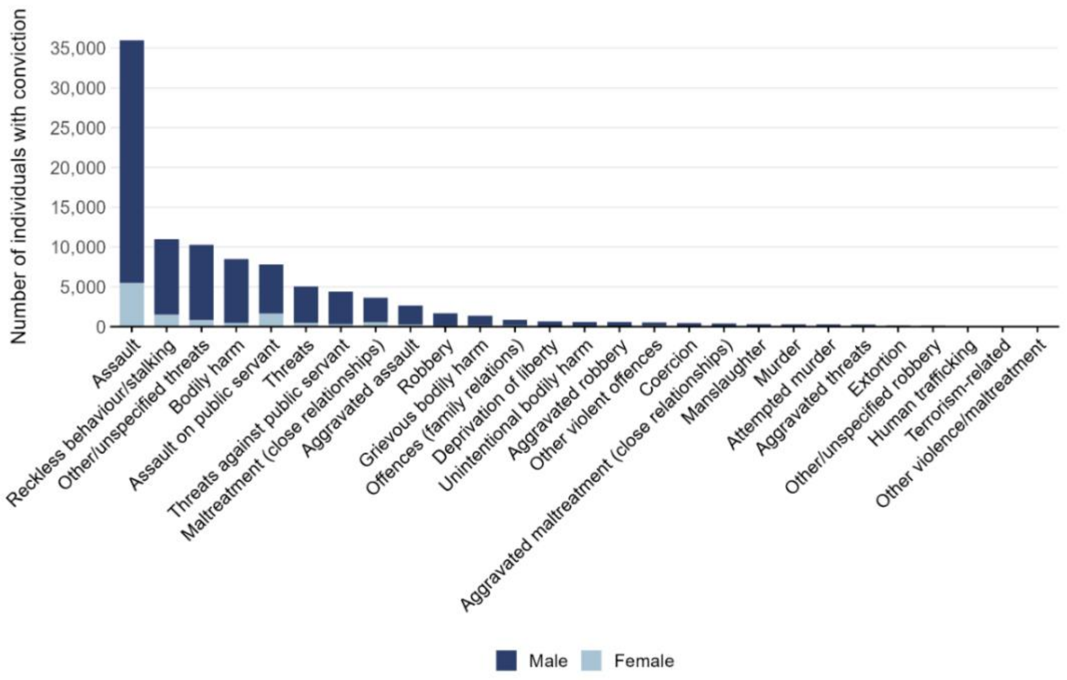

**Supplementary Figure 3a. Number of individuals convicted of each violent crime type, by presence of severe mental illness versus the remaining general population.**

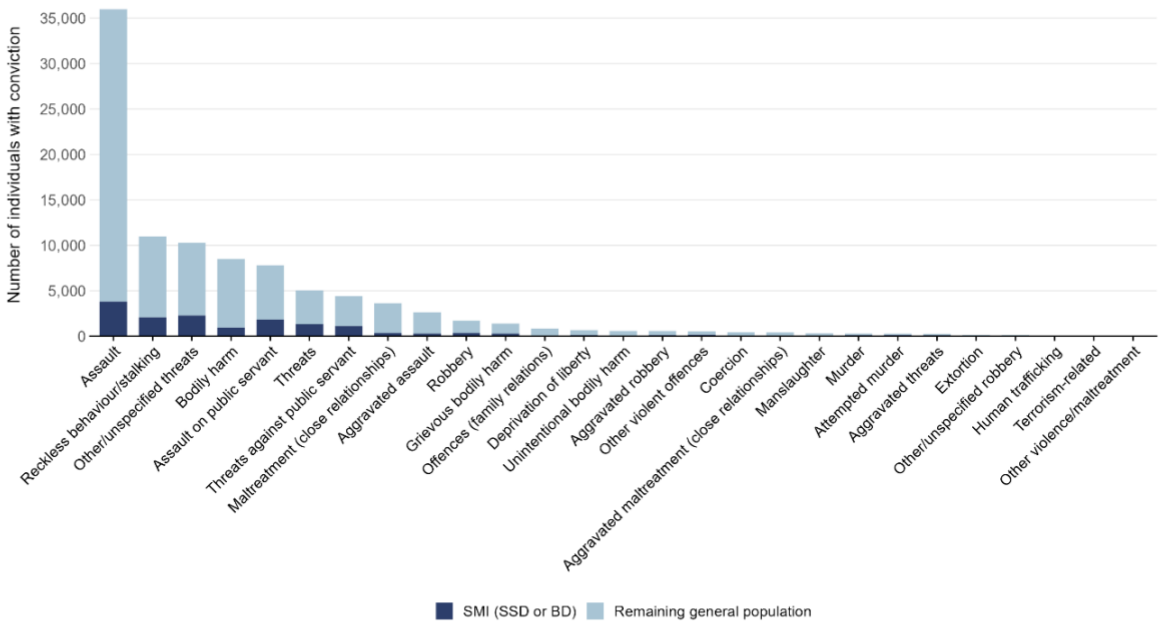

**Supplementary Figure 3b. Number of individuals convicted of each violent crime type, by presence of any mental disorder versus the remaining general population.**

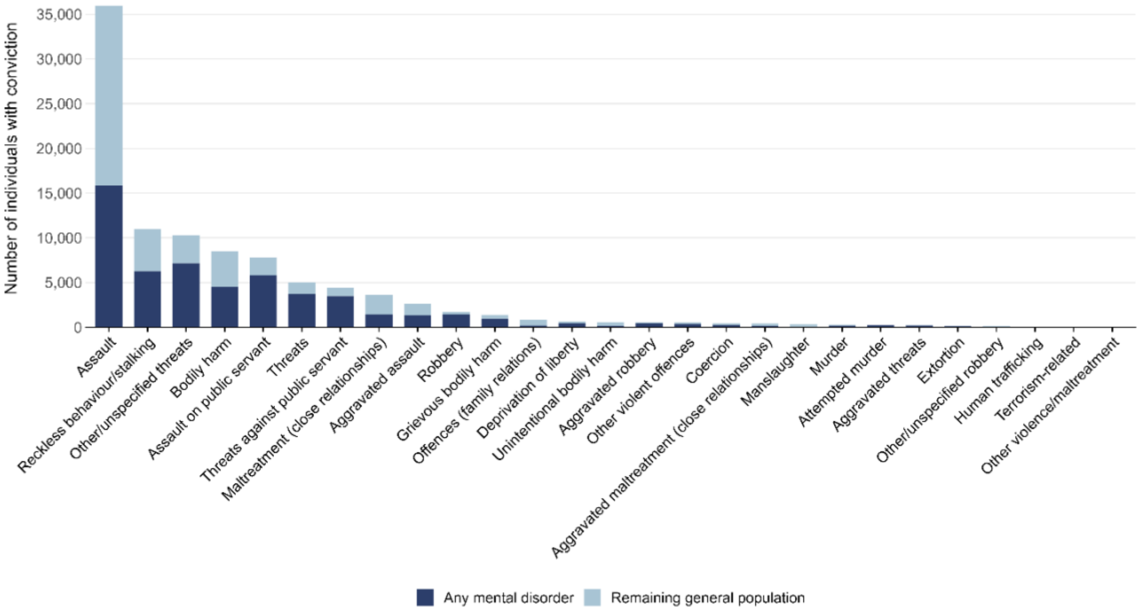

**Supplementary Table 3a: Relative risk of specific violent crime types associated with each mental disorder, adjusted for sex and age (Model 1).**

| | RR (95% CI) - Model 1: sex + age adjusted | | | | |
|---|---|---|---|---|---|
| **Violent crime type** | **SSD** | **BD** | **SUD** | **PD** | **ADHD** |
| Aggravated assault | 5·91 (5·15–6·78) | 2·82 (2·29–3·48) | 11·36 (10·51–12·28) | 10·86 (9·52–12·39) | 6·55 (5·92–7·24) |
| Aggravated maltreatment (close relationships) | 6·09 (4·22–8·78) | 2·58 (1·45–4·58) | 6·72 (5·42–8·34) | 14·34 (10·17–20·22) | 4·64 (3·31–6·50) |
| Aggravated robbery | 9·59 (7·53–12·20) | 3·79 (2·56–5·62) | 34·14 (28·39–41·06) | 23·36 (18·94–28·83) | 9·31 (7·69–11·27) |
| Aggravated threats | 31·08 (23·70–40·76) | 8·57 (5·62–13·05) | 27·06 (20·86–35·10) | 31·60 (23·33–42·82) | 9·13 (6·75–12·35) |
| Assault | 5·51 (5·31–5·72) | 3·35 (3·19–3·53) | 8·80 (8·62–8·98) | 8·10 (7·79–8·42) | 4·99 (4·84–5·14) |
| Assault on public servant | 15·78 (14·92–16·70) | 6·29 (5·79–6·83) | 31·46 (30·00–33·00) | 19·23 (18·10–20·43) | 9·26 (8·78–9·78) |
| Attempted murder | 41·79 (32·65–53·49) | 6·19 (3·94–9·73) | 22·05 (17·31–28·10) | 26·64 (19·61–36·18) | 6·84 (4·97–9·41) |
| Bodily harm | 5·74 (5·33–6·19) | 3·14 (2·81–3·51) | 11·67 (11·18–12·18) | 10·50 (9·77–11·29) | 5·99 (5·66–6·33) |
| Coercion | 7·19 (5·28–9·79) | 2·39 (1·38–4·15) | 19·13 (15·88–23·06) | 19·00 (14·56–24·80) | 11·97 (9·65–14·84) |
| Deprivation of liberty | 7·99 (6·27–10·18) | 4·81 (3·46–6·68) | 25·90 (22·06–30·40) | 21·91 (17·83–26·92) | 12·42 (10·44–14·78) |
| Extortion | 7·74 (4·62–12·98) | 4·05 (1·89–8·68) | 24·26 (17·49–33·63) | 18·60 (11·65–29·70) | 10·64 (7·27–15·58) |
| Grievous bodily harm | 13·16 (11·42–15·16) | 5·20 (4·18–6·47) | 26·45 (23·65–29·59) | 23·35 (20·31–26·85) | 9·18 (8·08–10·43) |
| Maltreatment (close relationships) | 5·71 (5·04–6·47) | 3·00 (2·51–3·58) | 7·67 (7·16–8·22) | 10·49 (9·25–11·89) | 4·75 (4·26–5·30) |
| Manslaughter | 1·67 (0·79–3·53) | 1·69 (0·75–3·79) | 5·13 (3·94–6·67) | 4·99 (2·72–9·15) | 3·52 (2·32–5·34) |
| Murder | 30·87 (24·01–39·69) | 5·51 (3·42–8·87) | 14·11 (11·19–17·78) | 27·07 (19·80–37·01) | 8·10 (5·92–11·09) |
| Offences (family relations) | 3·78 (2·74–5·21) | 2·46 (1·62–3·72) | 3·65 (3·04–4·37) | 4·06 (2·63–6·28) | 3·07 (2·26–4·17) |
| Other violent offences | 25·94 (21·41–31·43) | 6·31 (4·59–8·67) | 29·42 (24·64–35·12) | 28·70 (23·19–35·52) | 7·48 (5·96–9·39) |
| Other/unspecified robbery | 4·87 (2·47–9·59) | 1·31 (0·32–5·31) | 21·71 (15·14–31·13) | 23·01 (14·42–36·69) | 8·61 (5·65–13·13) |
| Other/unspecified threats | 14·75 (14·03–15·50) | 6·16 (5·71–6·65) | 23·32 (22·42–24·26) | 20·85 (19·76–22·01) | 9·01 (8·57–9·46) |
| Reckless behaviour/stalking | 12·03 (11·42–12·68) | 5·85 (5·43–6·30) | 14·28 (13·75–14·82) | 14·88 (14·01–15·80) | 7·11 (6·74–7·49) |
| Robbery | 14·13 (12·51–15·95) | 4·55 (3·69–5·60) | 60·29 (53·21–68·30) | 22·54 (19·95–25·47) | 9·26 (8·31–10·32) |
| Threats | 18·15 (16·98–19·41) | 7·61 (6·90–8·39) | 29·57 (27·89–31·34) | 21·70 (20·11–23·41) | 10·06 (9·39–10·77) |
| Threats against public servant | 17·09 (15·90–18·38) | 6·94 (6·22–7·74) | 39·08 (36·57–41·76) | 25·01 (23·15–27·00) | 10·85 (10·11–11·65) |
| Unintentional bodily harm | 1·45 (0·80–2·62) | 2·59 (1·60–4·20) | 3·68 (2·98–4·55) | 2·68 (1·47–4·87) | 2·77 (1·95–3·92) |

*Poisson regression with robust standard errors (no time offset). Outcome: any violent crime conviction (binary yes/no). Reference group: remaining general population (all individuals without the disorder of interest). Model 1: adjusted for sex and age. Model 2: additionally adjusted for all other mental disorders (SSD, BD, SUD, PD, ADHD). RR = relative risk; CI = confidence interval. SSD = schizophrenia spectrum disorders (F20–F29); BD = bipolar disorder (F30–F31); SUD = substance use disorders (F10–F19); PD = common personality disorders (F60·0, F60·1, F60·2, F60·3, F60·4); ADHD = hyperkinetic disorders (F90).*

**Supplementary Table 3b: Relative risk of specific violent crime types associated with each mental disorder, fully adjusted for co-occurring mental disorders (Model 2).**

| | RR (95% CI) - Model 2: fully adjusted | | | | |
|---|---|---|---|---|---|
| **Violent crime type** | **SSD** | **BD** | **SUD** | **PD** | **ADHD** |
| Aggravated assault | 1·53 (1·32–1·77) | 0·76 (0·61–0·94) | 8·23 (7·50–9·03) | 2·39 (2·06–2·77) | 2·30 (2·06–2·57) |
| Aggravated maltreatment (close relationships) | 2·08 (1·38–3·12) | 0·80 (0·44–1·46) | 4·62 (3·51–6·08) | 4·59 (3·05–6·90) | 1·74 (1·18–2·56) |
| Aggravated robbery | 1·49 (1·16–1·93) | 0·66 (0·44–0·98) | 23·60 (19·22–28·96) | 3·47 (2·75–4·39) | 2·04 (1·66–2·50) |
| Aggravated threats | 5·91 (4·12–8·48) | 1·26 (0·80–1·97) | 12·99 (9·17–18·39) | 3·72 (2·58–5·38) | 1·83 (1·30–2·57) |
| Assault | 1·70 (1·63–1·77) | 1·08 (1·02–1·14) | 6·71 (6·55–6·88) | 2·02 (1·93–2·10) | 1·93 (1·86–1·99) |
| Assault on public servant | 2·79 (2·62–2·97) | 1·08 (0·99–1·18) | 20·56 (19·45–21·74) | 2·47 (2·31–2·64) | 2·03 (1·92–2·15) |
| Attempted murder | 10·57 (7·33–15·24) | 0·90 (0·55–1·47) | 9·56 (6·75–13·53) | 3·16 (2·18–4·57) | 1·45 (1·03–2·04) |
| Bodily harm | 1·45 (1·33–1·57) | 0·85 (0·76–0·96) | 8·74 (8·31–9·19) | 2·30 (2·13–2·49) | 2·06 (1·94–2·19) |
| Coercion | 1·40 (1·01–1·94) | 0·47 (0·27–0·82) | 12·11 (9·73–15·07) | 3·19 (2·37–4·30) | 3·38 (2·65–4·32) |
| Deprivation of liberty | 1·34 (1·04–1·72) | 0·88 (0·63–1·23) | 16·39 (13·57–19·78) | 3·25 (2·57–4·10) | 3·02 (2·48–3·67) |
| Extortion | 1·38 (0·80–2·39) | 0·78 (0·35–1·75) | 16·74 (11·30–24·81) | 2·87 (1·64–5·02) | 2·63 (1·69–4·10) |
| Grievous bodily harm | 2·30 (1·97–2·69) | 0·91 (0·73–1·14) | 16·98 (14·92–19·33) | 3·41 (2·91–4·00) | 2·06 (1·79–2·36) |
| Maltreatment (close relationships) | 1·91 (1·66–2·20) | 0·97 (0·80–1·17) | 5·72 (5·26–6·21) | 2·91 (2·51–3·36) | 1·81 (1·60–2·04) |
| Manslaughter | 0·70 (0·33–1·51) | 0·79 (0·34–1·81) | 4·59 (3·44–6·13) | 1·96 (1·01–3·80) | 1·79 (1·14–2·81) |
| Murder | 9·07 (6·22–13·24) | 0·91 (0·55–1·51) | 5·82 (4·14–8·19) | 3·87 (2·62–5·71) | 2·03 (1·45–2·85) |
| Offences (family relations) | 2·14 (1·51–3·02) | 1·28 (0·83–1·97) | 2·96 (2·40–3·66) | 1·60 (0·99–2·60) | 1·75 (1·26–2·44) |
| Other violent offences | 4·91 (3·92–6·16) | 0·95 (0·68–1·33) | 16·69 (13·45–20·72) | 3·71 (2·91–4·73) | 1·47 (1·16–1·87) |
| Other/unspecified robbery | 0·85 (0·42–1·69) | 0·26 (0·06–1·04) | 15·74 (10·38–23·86) | 4·51 (2·54–7·99) | 2·14 (1·31–3·49) |
| Other/unspecified threats | 2·89 (2·74–3·06) | 1·13 (1·04–1·23) | 15·23 (14·53–15·97) | 2·90 (2·73–3·09) | 2·03 (1·93–2·15) |
| Reckless behaviour/stalking | 3·02 (2·84–3·21) | 1·35 (1·24–1·46) | 9·42 (8·99–9·86) | 2·51 (2·34–2·69) | 1·99 (1·88–2·11) |
| Robbery | 2·02 (1·78–2·29) | 0·71 (0·57–0·87) | 42·60 (37·27–48·71) | 2·79 (2·44–3·18) | 1·79 (1·60–2·00) |
| Threats | 3·29 (3·05–3·55) | 1·28 (1·16–1·42) | 18·41 (17·19–19·72) | 2·63 (2·42–2·87) | 2·12 (1·97–2·28) |
| Threats against public servant | 2·73 (2·52–2·95) | 1·09 (0·98–1·22) | 24·98 (23·16–26·94) | 2·97 (2·73–3·23) | 2·11 (1·96–2·28) |
| Unintentional bodily harm | 0·74 (0·40–1·36) | 1·56 (0·93–2·60) | 3·34 (2·66–4·20) | 1·18 (0·62–2·22) | 1·66 (1·16–2·40) |

*Poisson regression with robust standard errors. Outcome: any violent crime conviction (binary yes/no). Reference group: remaining general population (all individuals without the disorder of interest). Model 1: adjusted for sex and age. Model 2: additionally adjusted for all other mental disorders (SSD, BD, SUD, PD, ADHD). RR = relative risk; CI = confidence interval. SSD = schizophrenia spectrum disorders (F20–F29); BD = bipolar disorder (F30–F31); SUD = substance use disorders (F10–F19); PD = common personality disorders (F60·0, F60·1, F60·2, F60·3, F60·4); ADHD = hyperkinetic disorders (F90).*
